# A Pilot Teaching Kitchen Project Increases Confidence for Dietary Interventions Aimed at Prevention of Kidney Stones in Known Stone Formers

**DOI:** 10.1101/2025.07.31.25330593

**Authors:** Ellie Mehrara, Ava Mousavi, Sapna Thaker, Karan Thaker, Kyle Zuniga, Shelby Yaceczko, Lynn Stothers, Kymora B Scotland

## Abstract

**Introduction:** Patterns of dietary intake are known risk factors for both kidney stone formation and recurrence.^1,2^ Dietary recommendations such as the Dietary Approaches To Stop Hypertension (DASH) diet can reduce risk,^3,4^ but patients may lack knowledge or a practical understanding on implementing these diet patterns. Virtual teaching kitchens have proven effective in improving dietary practices in other populations, but have not been applied to patients with kidney stones.^5,6^ This study assessed the effectiveness of virtual teaching kitchen sessions in enhancing dietary confidence and practices for kidney stone prevention.

**Methods:** Eligible adults, aged 18 years and older, with a history of recurrent kidney stones were recruited to participate in one of two virtual teaching kitchen sessions. Pre-intervention surveys were administered to participants assessing individual cooking habits and dietary confidence. Post-intervention surveys assessed changes in cooking attitudes, dietary confidence, and program satisfaction.

**Results:** Forty-six participants completed both pre- and post-intervention surveys. Results showed significant improvements in enjoyment of trying new recipes, reduced frustration with cooking, and perceptions of home-cooked meals as both affordable and healthy. Participants also reported finding cooking less tiring and increased confidence in adopting key dietary practices, including reducing added sugar intake, choosing lean proteins, using spices instead of salt, consuming more fruits and vegetables, and controlling portion sizes (all p<0.05).

**Conclusions:** The virtual teaching kitchen intervention significantly improved participants’ confidence and attitudes toward adopting dietary practices essential for kidney stone prevention, highlighting its promise as an educational tool for patients.

## Introduction

Kidney stones affect about 10% of the population in the United States, with prevalence increasing every year.^1^ These stones represent one of the most costly urologic conditions, with costs exceeding $10 billion each year.^1^ Kidney stones are associated with increased risk of comorbidities including metabolic syndrome, cardiovascular disease, and chronic kidney disease.^1,2^ Calcium oxalate stones are the most common type of kidney stones, and are associated with a high rate of recurrence in patients, mainly due to a lack of appropriate management of the factors that increase the risk of stone formation.^3^

Poor nutrition and highly processed diet patterns have been identified as significant contributors to kidney stone formation and recurrence, primarily due to their impact on urine chemistry.^1^ Specifically, certain dietary patterns can increase the risk of calcium oxalate crystallization in the urine. Under regular digestive conditions, calcium and oxalate bind in the intestines and are excreted in the stool. However, excess sodium in the diet disrupts this balance by promoting increased urinary calcium excretion. Similarly, excessive consumption of oxalate-rich foods, such as spinach, berries, and chocolate, raise urinary oxalate levels when not adequately bound by intestinal calcium, thereby facilitating stone formation.^2^ Dietary recommendations for kidney stone prevention directly address these issues. Increased fluid intake is encouraged to dilute urine and prevent mineral supersaturation, while reducing sodium intake helps lower urinary calcium excretion. Additionally, moderating consumption of oxalate-rich foods can help reduce oxalate levels in the urine. A balanced calcium intake is also recommended to ensure that dietary calcium can bind oxalate in the intestines. Separately, maintaining a healthy body weight has been associated with a reduced risk of kidney stone formation, as obesity is a known risk factor. Incorporating more vegetables and fiber into the diet can support improved digestion and reduce the absorption of stone-forming substances.^1^ However, these dietary changes can be challenging to implement without specific guidance and support.

Prior studies have reported that the dietary approach referred to as the Dietary Approaches to Stop Hypertension (DASH) diet has been associated with a significant decrease in the risk of kidney stone formation and recurrence.^1,4^ This dietary pattern is characterized by a high intake of fruits and vegetables, nuts, legumes, whole grains, and low fat dairy and a limited intake of added sugar, processed meat, and sodium. Taylor et al used a combined 50 year follow up to evaluate the relationship between a DASH style diet and incident kidney stones in over 240,000 participants. Participants with higher adherence to the diet (higher DASH score) showed lower relative risks for kidney stones than those with lower adherence.^4^ Moreover, Maddahi et al evaluated 264 participants with a history of nephrolithiasis. They found that higher adherence to a DASH style diet was associated with lower risk of hypercreatinemia, hypocitraturia, and hypercalciuria, all of which are risks for the formation of kidney stones.^3^ These studies both showed that consumption of a DASH style diet was associated with a decrease in the risk of incident kidney stones as well as a beneficial change in urine chemistry.^3,4^ Because of this, clinicians can recommend the DASH diet as a dietary approach to improve modifiable risk factors of kidney stone formation to patients.

Teaching kitchens serve as experiential learning environments designed to impart culinary skills, nutritional knowledge, and evidence-based cooking practices, with the goal of fostering healthier dietary behaviors and improved eating patterns.^5^ These kitchens have proven to be valuable tools for the improvement of eating habits and certain biomarkers in individuals with type 2 diabetes and perimenopausal women.^5,6^ A study conducted by Sommer et al. evaluated the effectiveness of a virtual teaching kitchen to increase confidence and consumption of a healthy diet in perimenopausal women. After instructing 609 participants, they found that virtual teaching kitchens were effective in improving cooking confidence and dietary habits. Patients reported an increase in confidence in their ability to eat healthy and cook for themselves, increased knowledge of healthy recipes, and beneficial modifications to their diets.^5,6^ Moreover, virtual formats of these teaching kitchens have been proven to be effective and efficient at teaching patients proper cooking techniques and educating them on implementing dietary changes into their everyday lives.^5^ Virtual teaching kitchens are less costly, allow for an increased number of participants, provide more flexibility in the times that they can take place, and can be more interactive. Given these benefits, this study aimed to determine if a virtual teaching kitchen is effective at educating patients with kidney stones on nutrition recommendations and evidence based dietary patterns to prevent kidney stone recurrence and formation. We hypothesized that a virtual teaching kitchen would be effective in educating patients with kidney stones on evidence-based stone prevention diets and cooking techniques, thereby increasing confidence in adopting healthier dietary practices.

## Materials and Methods

### Virtual Teaching Kitchen Session Preparation

This study involved two virtual teaching kitchen sessions conducted via HIPAA-compliant Zoom. Men and women aged 18 and above with working access to Zoom and previous incidents of kidney stones and kidney stone recurrences were eligible to participate in the virtual teaching kitchen. Participants previously seen for kidney stone disease at a tertiary care endourology clinic were recruited via email. Following informed consent, subjects received reminder emails with session details, preparation guidance, recipes, and required ingredients.

Prior to each virtual teaching kitchen session, participants were asked to complete a seven-question pre-intervention survey administered via Qualtrics. Survey questions were adopted from a modified version of the Cooking Attitude Survey and included items assessing participants’ cooking habits and current use of a stone prevention diet.^7^ Responses were collected using a 5-point Likert scale, where 1 represented ‘strongly disagree,’ 2 ‘disagree,’ 3 ‘neither agree nor disagree,’ 4 ‘agree,’ and 5 ‘strongly agree.’

### Virtual Teaching Kitchen Session

Participants joined each virtual teaching kitchen via a password-protected Zoom call. Each session began with a brief lecture providing an overview of a kidney stone prevention diet, implementation strategies, and guidance based on content from the “Living Healthy: Fight Kidney Stones With Food Cookbook, a freely available online cookbook published by the American Urological Association.

The lecture addressed specific dietary recommendations essential for kidney stone prevention, emphasizing the importance of fluid intake, daily servings of fruits and vegetables, consideration of oxalate levels, reduced meat consumption, calcium intake, and sodium limitation.

Following the lecture, participants were guided through the preparation and cooking of four recipes: The first session taught participants how to make a “Fresh Blueberry and Lemon Smoothie” and “Oven Omelets”. The second session taught participants how to make a “Peachy Strawberry Slush Drink” and “Robust Chicken Curry” (Fig. 1). Participants were given the recipes and instructions in advance, enabling them to follow along from their own kitchens or just observe. The instructors, who included two urologists and a registered dietitian, advised participants on each step of the recipes, proper techniques, and tips to customize the food to their liking. Throughout both recipe demonstrations, the instructors encouraged questions and interaction, fostering an engaging and supportive learning environment. Participants were given real-time feedback on their techniques and modifications, ensuring they felt confident in replicating the recipes independently. Both virtual sessions followed an identical structure and curriculum.

**Figure 1.**
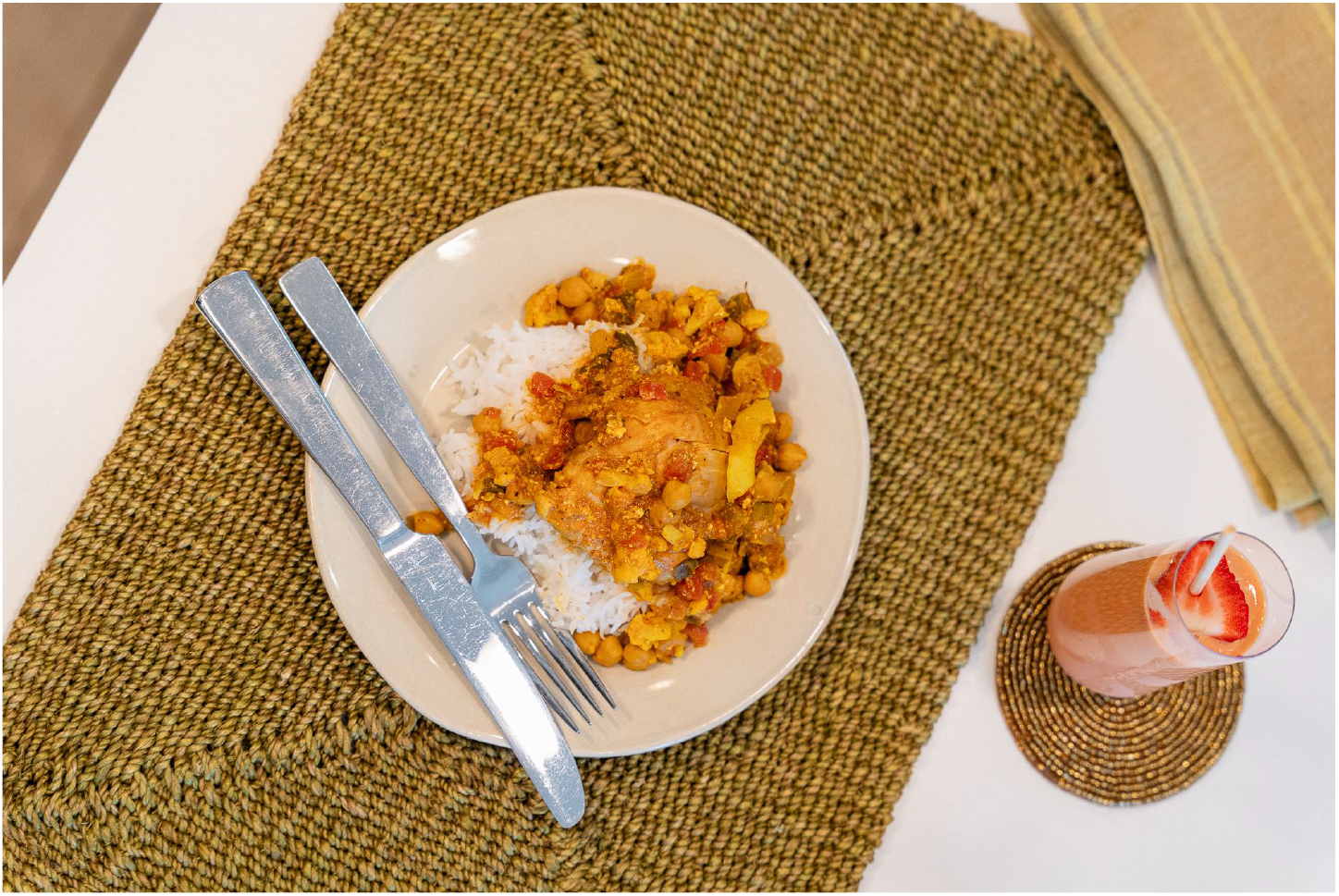
“Robust Chicken Curry” and “Peachy Strawberry Slush Drink” Prepared During the Virtual Teaching Kitchen Session.

### Post-Session Surveys

After the virtual teaching kitchen session, participants completed a 12-question survey, which was administered again through Qualtrics. This survey included questions from modified versions of the Cooking Attitude Survey and the Eating Habits Confidence Survey, to assess participants’ feelings about preparing their own meals after the lesson and their satisfaction with the training.^5,7^ The same 5-point Likert scale employed in the pre-intervention survey was also used in the post-intervention survey. Survey responses were collected and analyzed separately and in aggregate (Fig. 2).

**Figure 2.**
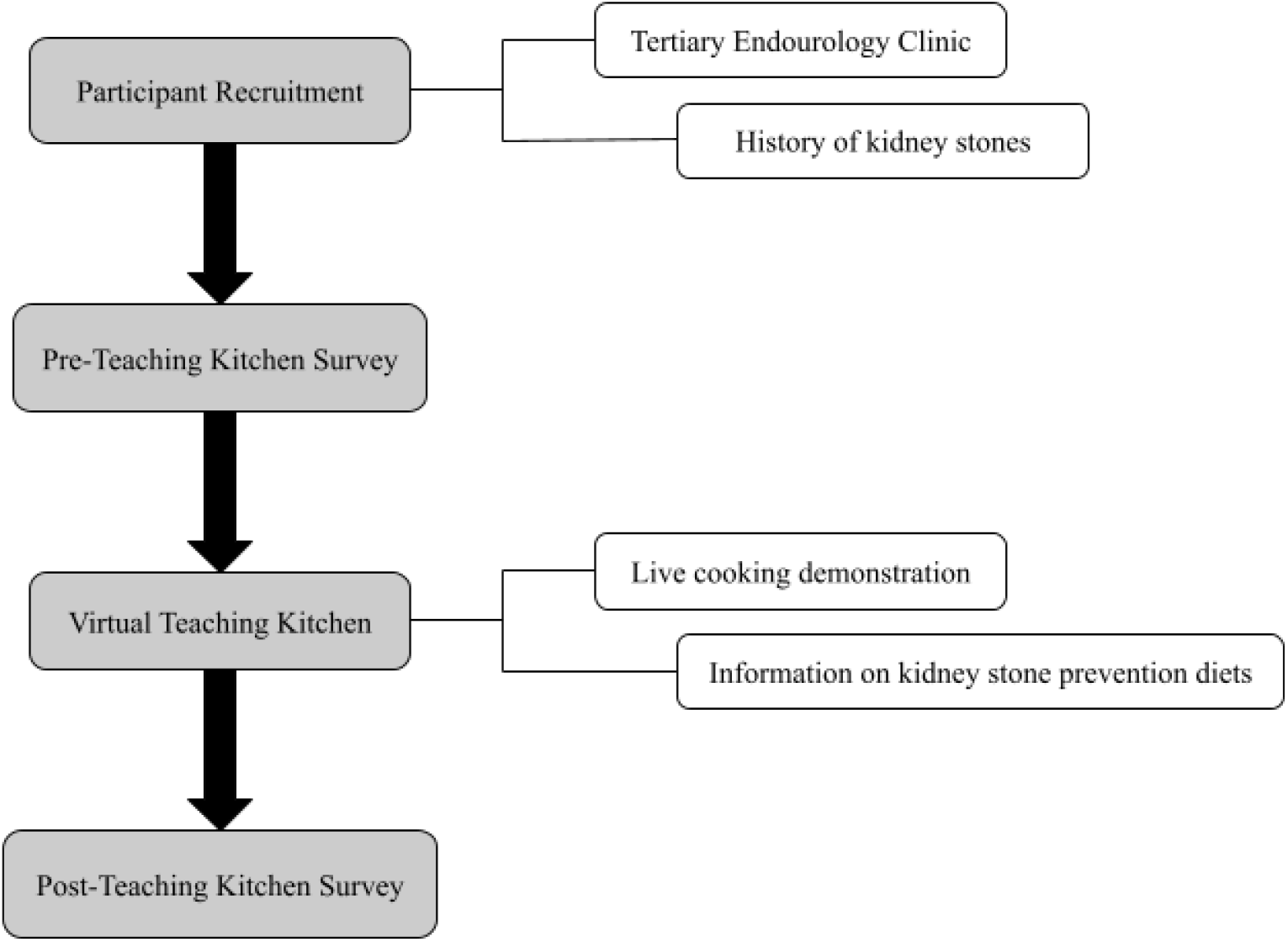
Flowchart demonstrating the multi-step procedure utilized in evaluating the effectiveness of a virtual teaching kitchen for kidney stone patients.

### Statistical Considerations

The primary outcome of this study was participant satisfaction with the virtual teaching kitchen and their confidence in preparing their own meals before and after the session. To assess confidence levels, a Likert scale was employed with pre- and post-session data being summarized into one binary variable (improved vs. not-improved). Data from both sessions were pooled for analysis. A single-group design was used to test whether the proportion is different from 0.1. The comparison was made using a two-sided, one-sample exact test, with a significance level of 5% and power of 80%. Based on preliminary calculations, we determined that a sample size of 22 participants was required to achieve statistical significance. However, we chose to double this number to account for potential dropout on the day of the intervention and due to high patient interest.

## Results

Two virtual teaching kitchens were conducted as part of this study. In the first session, 25 participants attended; of these, 16 completed both the pre- and post-intervention surveys. Two additional responses were excluded due to duplicate pre-teaching kitchen survey responses, resulting in 14 valid paired responses. In the second session, 35 participants attended; of these, 34 completed both the pre- and post-intervention surveys. Two responses were excluded - one due to a duplicate pre-teaching kitchen survey response and one due to a duplicate post-teaching kitchen survey response - yielding 32 valid paired responses. In total, 46 participants across both sessions completed both the pre- and post-surveys (Table 1).

**Table 1:** Demographic characteristics of virtual teaching kitchen participants (n = 46)

| Teaching Kitchen Participant Demographics |  |
| --- | --- |
| Male Participants | 41.3% |
| Female Participants | 58.7% |
| Mean Participant Age | 61.1 |

After participating in the virtual teaching kitchen events, there was a statistically significant improvement in several aspects of participants’ attitudes toward cooking (Table 2). Notably, more participants reported enjoying trying new recipes (p = 0.0030) and feeling that cooking was less frustrating (p = 0.0097). There was also a significant increase in agreement that meals made at home are affordable (p = 0.0281) and that home cooking supports healthier eating (p = 0.0281). Additionally, participants were significantly less likely to find cooking tiring following the intervention (p = 0.0008).

**Table 2:** Pre-to Post-Intervention Changes in Eating and Cooking Confidence.

| **Views About Cooking (1 = Strongly Disagree, 2 = Disagree, 3 Neither Agree Nor Disagree, 4 = Agree, 5 = Strongly Agree)** | **Proportion Improved After Teaching Kitchen** | **Proportion Not Improved After Teaching Kitchen** | **p Value** |
| --- | --- | --- | --- |
| I do NOT like to cook because it takes too much time. | 0.196 | 0.804 | 0.0727 |
| Meals made at home are affordable. | 0.217 | 0.783 | 0.0281* |
| Cooking is frustrating. | 0.239 | 0.761 | 0.0097* |
| I like trying new recipes. | 0.261 | 0.739 | 0.0030* |
| It is too much work to cook. | 0.196 | 0.804 | 0.0727 |
| Making meals at home helps me to eat more healthfully. | 0.217 | 0.783 | 0.0281* |
| I find cooking tiring. | 0.283 | 0.717 | 0.0008* |

The virtual teaching kitchen also had a significant impact on participants’ confidence in adopting healthier dietary practices essential for kidney stone prevention (Table 3). Participants showed a significant increase in confidence in their ability to consume foods and beverages with less added sugar (p<0.001), choose lean meats instead of red meat (p<0.001), use spices to avoid adding salt at the table (p<0.001), eat at least four to five servings of fruits and vegetables per day (p<0.001), and avoid overly large portions (p<0.001).

**Table 3:**
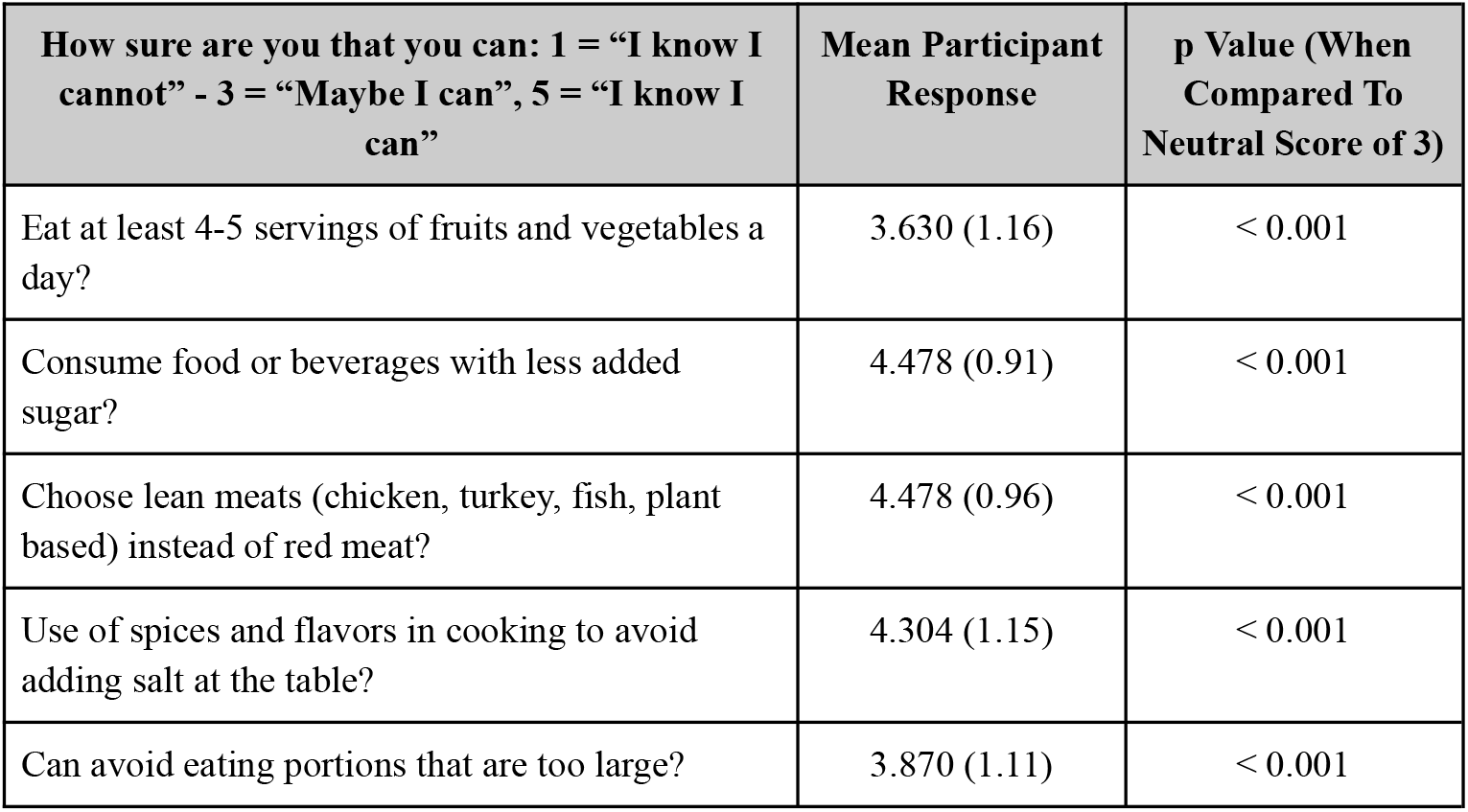
Eating Habits Confidence of Participants Post Teaching Kitchen.

## Discussion

The results of our study indicate that the virtual teaching kitchen was effective in improving participants’ attitudes towards cooking and their confidence in implementing dietary changes necessary for kidney stone prevention. The significant improvement in participants’ willingness to try new recipes and view cooking as a less tiring activity suggests that the teaching kitchen positively influenced their perceptions and enjoyment of cooking, which is essential for sustainable dietary changes.

A notable outcome was the significant increase in participants’ confidence to adopt healthier dietary practices. Participants reported greater confidence in reducing added sugar intake, which indicates a greater awareness and willingness to make healthier choices regarding added sugar consumption. They also showed increased confidence in choosing lean meats over red meat. Furthermore, participants were more confident in using spices, instead of adding salt at the table, reflecting a positive shift toward flavoring foods in a manner without increasing sodium intake. Additionally, survey results reported more confidence in consuming the recommended daily servings of fruits and vegetables and avoiding overly large portions of overall food intake, which are critical for maintaining a healthy weight and reducing kidney stone risk. These dietary changes align with the recommendations for preventing kidney stones and demonstrate the program’s effectiveness in imparting practical, actionable knowledge.^8^

Our study adds to the growing body of evidence supporting the effectiveness of virtual teaching kitchens. Previous research has shown that teaching kitchens can improve dietary habits and certain health biomarkers in various populations, including individuals with type 2 diabetes and perimenopausal women.^5,6^ Our findings are consistent with these studies, demonstrating that virtual teaching kitchens can effectively enhance cooking confidence and promote healthier eating patterns among kidney stone patients.

An important aspect of the study design was the deliberate inclusion of urologists, in addition to a nutritionist, to model cooking and demonstrate healthy dietary practices. Notably, one of the urologists had an established relationship with the participants through prior clinical care, which likely enhanced participants’ receptiveness to the dietary guidance and made the experience more personal. This aligns with social learning theory, which supports the idea that using a trusted figure practicing the behavior change can be motivational. This is an extension of findings from heart health and diabetes management which support the use of multidisciplinary teams (including physicians and dieticians) in delivering effective dietary education.^9,10^ Additionally, hosting the teaching kitchen in a home kitchen setting, rather than a clinical environment, provided a more practical and relatable setting that encouraged participants to apply the cooking techniques in their own lives.

The virtual format of the teaching kitchen offers several advantages, including lower costs, increased accessibility, and flexibility in scheduling. These benefits are particularly important given the busy lifestyles and varied schedules of participants, allowing them to integrate learning into their daily routines more easily. The interactive nature of the virtual sessions, with real-time feedback and opportunities for participant engagement, likely contributed to the program’s success.

However, several limitations should be considered when interpreting these results. The findings were limited to a single institution and may have limited generalizability to broader populations. While our sample included a range of demographic backgrounds, all participants had sufficient English language proficiency to follow the virtual teaching kitchen sessions. Future studies should aim to incorporate participants with limited English proficiency to ensure the intervention is accessible and effective across diverse populations. Additionally, the self-selected nature of the participants may introduce selection bias, as individuals who chose to participate may have had a pre-existing interest in cooking or dietary health. Although the study did not include a separate control group, the pre- and post-survey design allows each participant to serve as their own control, strengthening the internal validity of the findings. Nonetheless, future studies would benefit from including a control group to more rigorously isolate the intervention’s effects. In addition, conducting follow-up surveys several months post-intervention could help assess the long-term sustainability of the dietary changes and confidence gains observed immediately after the program.

Finally, while this study focused on participant confidence and attitudes, it did not directly address clinical outcomes such as changes in urinary stone risk or recurrence rates. Future studies should incorporate objective measures, including biochemical assessments of urine composition and stone risk factors, to evaluate the direct impact of virtual teaching kitchens on kidney stone prevention.

## Conclusions

The virtual teaching kitchen significantly improved participants’ attitudes towards cooking and their confidence in adopting dietary practices essential for kidney stone prevention. The findings suggest that virtual teaching kitchens are a promising tool for educating patients with kidney stones on evidence-based diet patterns and cooking techniques for kidney stone prevention. Future research with additional objective measures of dietary intake and behavior is needed to further validate these findings and assess the long-term impact of such interventions.

## Data Availability

All data produced in the present study are available from the corresponding author upon reasonable request.

## Notes

### Competing Interest Statement

The authors have declared no competing interest.

### Funding Statement

This study did not receive any funding.

### Author Declarations

The Institutional Review Board of the University of California, Los Angeles gave ethical approval for this work.

